# Pediatric Nirmatrelvir/Ritonavir Prescribing Patterns During the COVID-19 Pandemic

**DOI:** 10.1101/2022.12.23.22283868

**Authors:** Seuli Bose-Brill, Kathryn Hirabayashi, Nathan M Pajor, Suchitra Rao, Asuncion Mejias, Ravi Jhaveri, Christopher B. Forrest, Charles Bailey, Dimitri A. Christakis, Deepika Thacker, Patrick C. Hanley, Payal B. Patel, Jonathan D. Cogen, Jason P. Block, Priya Prahalad, Vitaly Lorman, Grace M. Lee, the RECOVER consortium

## Abstract

**Objective:** This study was conducted to identify rates of pediatric nirmatrelvir/ritonavir (Paxlovid) prescriptions overall and by patient characteristics.

**Methods:** Patients up to 23 years old with a clinical encounter and a nirmatrelvir/ritonavir (Paxlovid, n/r) prescription in a PEDSnet-affiliated institution between December 1, 2021 and September 14, 2022 were identified using electronic health record (EHR) data.

**Results:** Of the 1,496,621 patients with clinical encounters during the study period, 920 received a nirmatrelvir/ritonavir prescription (mean age 17.2 years; SD 2.76 years). 40% (367/920) of prescriptions were provided to individuals aged 18-23, and 91% (838/920) of prescriptions occurred after April 1, 2022. The majority of patients (70%; 648/920) had received at least one COVID-19 vaccine dose at least 28 days before nirmatrelvir/ritonavir prescription. Only 40% (371/920) of individuals had documented COVID-19 within the 0 to 6 days prior to receiving a nirmatrelvir/ritonavir prescription. 53% (485/920) had no documented COVID-19 infection in the EHR. Among nirmatrelvir/ritonavir prescription recipients, 64% (586/920) had chronic or complex chronic disease and 9% (80/920) had malignant disease. 38/920 (4.5%) were hospitalized within 30 days of receiving nirmatrelvir/ritonavir.

**Conclusion:** Clinicians prescribe nirmatrelvir/ritonavir infrequently to children. While individuals receiving nirmatrelvir/ritonavir generally have significant chronic disease burden, a majority are receiving nirmatrelvir/ritonavir prescriptions without an EHR-recorded COVID-19 positive test or diagnosis. Development and implementation of concerted pediatric nirmatrelvir/ritonavir prescribing workflows can help better capture COVID-19 presentation, response, and adverse events at the population level.

## INTRODUCTION

The United States (U.S.) Food and Drug Administration (FDA) issued an Emergency Use Authorization (EUA) in December, 2021 allowing nirmatrelvir/ritonavir (Paxlovid) administration to children ≥ 12 years for treatment of mild-to-moderate COVID-19, if at risk of progression to severe disease.^1^

The eight-fold increased risk of COVID-19 related hospitalization among children with chronic disease compared to those without ^2,3^ underscores the potential utility of pediatric nirmatrelvir/ritonavir use.^4^ Yet, extrapolation of adult data and recommendation primarily informs pediatric nirmatrelvir/ritonavir treatment guidance,^5^ as pediatric data on feasibility, safety, efficacy, and toxicity remain limited. ^6, 7^ In this environment, evaluation of national pediatric nirmatrelvir/ritonavir prescribing data can help guide clinical decision-making and observational study design.^8^ This study leverages data from PEDSnet, a national pediatric learning health system, to characterize clinical features associated with nirmatrelvir/ritonavir prescribing in children.^9^

## METHODS

This 10-month retrospective cohort study of electronic health record (EHR) data from 8 PEDSnet health systems, collectively providing services to 3.3% of the nation’s children (2.4 million patients) annually,^9,2^ includes patients aged 0-23 years with any type of visit from December 1, 2021 to September 29, 2022.^1^ The Children’s Hospital of Philadelphia’s institutional review board designated this study as not human subjects research.

Study cohort patients prescribed nirmatrelvir/ritonavir during study period were identified, along with COVID-19 status and hospitalization occurring within 1 day prior to 30 days following prescription. The Pediatric Medical Complexity Algorithm (PMCA) Version 2.0 was used to categorize children based on chronic disease comorbidity burden.^10^

Descriptive statistics were calculated on the week 129 PEDSnet Researching COVID to Enhance Recovery (RECOVER) database (data loading cutoff: 9/29/2022) using R version 4.2.0 (2022-04-2022).

## RESULTS

Of the 1,496,621 individuals aged 0-23 years (mean: 7.3; SD: 2.76) in the PEDSnet RECOVER database, 920 received a nirmatrelvir/ritonavir prescription (Table 1), the majority [91% (838/920)] occurring after April 1, 2022 (Figure 1). Few prescriptions (<5) were dispensed outside of the FDA’s EUA age specifications, to children aged 10-11 years. ^1^ The majority of nirmatrelvir/ritonavir prescription recipients were non-Hispanic White, and female (Table 1), and ∼60% (∼553/920) were ≤ 17 years old. The majority of recipients, 70% (648/920), had received at least 1 COVID-19 vaccination at least 28 days prior to nirmatrelvir/ritonavir prescription. Notably, 47% (435/920) of nirmatrelvir/ritonavir prescription recipients had EHR documentation of COVID-19, while 53% (485/920) of recipients had none. 41% (374/920) of recipients had EHR COVID-19 documentation in the 0-6 days before prescription. Among nirmatrelvir/ritonavir recipients with EHR documentation of COVID-19, 40% (172/435) had only an outpatient COVID-19 diagnosis code, 8% (35/435) had only a positive polymerase chain reaction test, and 52% (228/435) had multiple positive COVID-19 tests and/ or outpatient diagnosis codes.

**Table 1.**
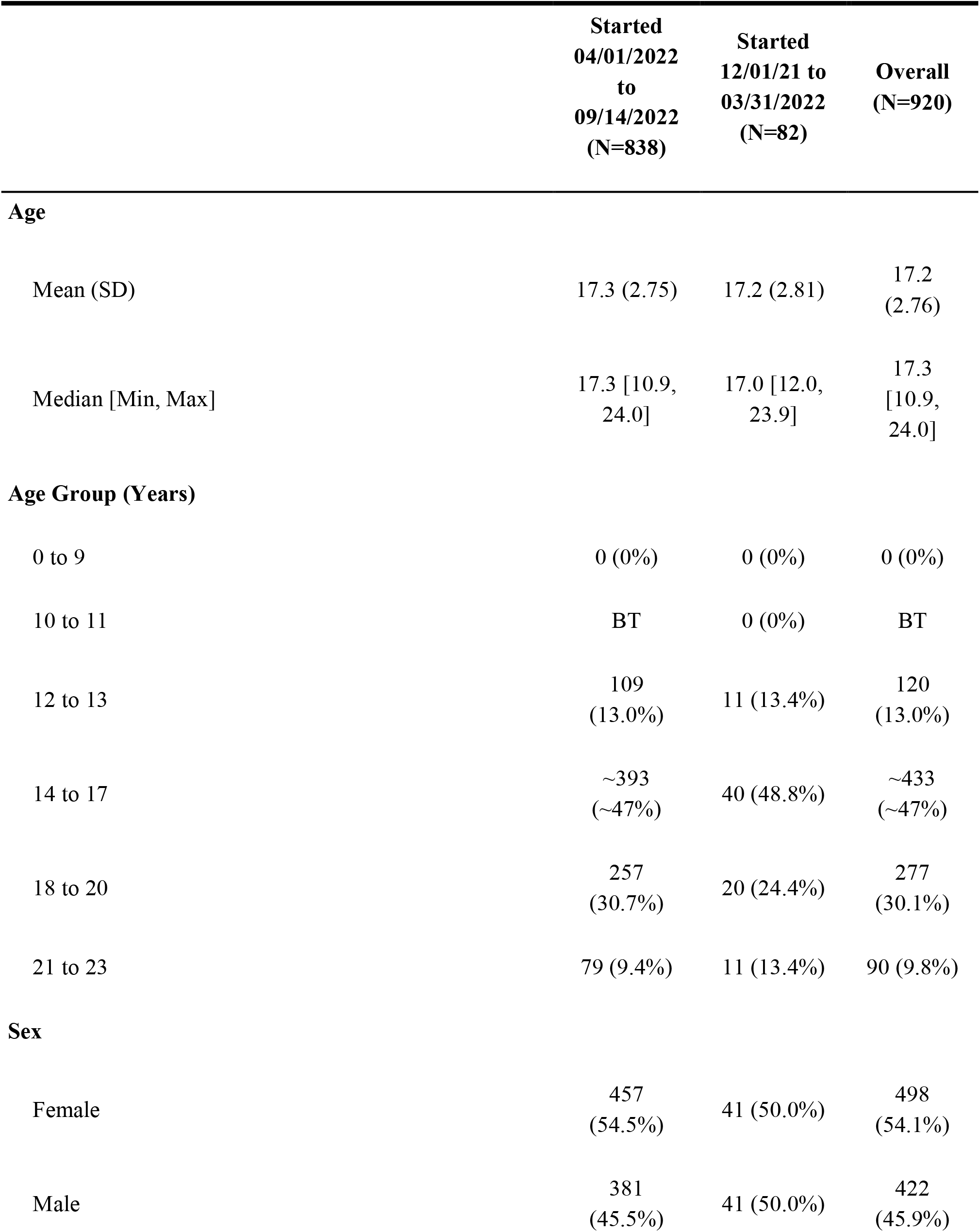
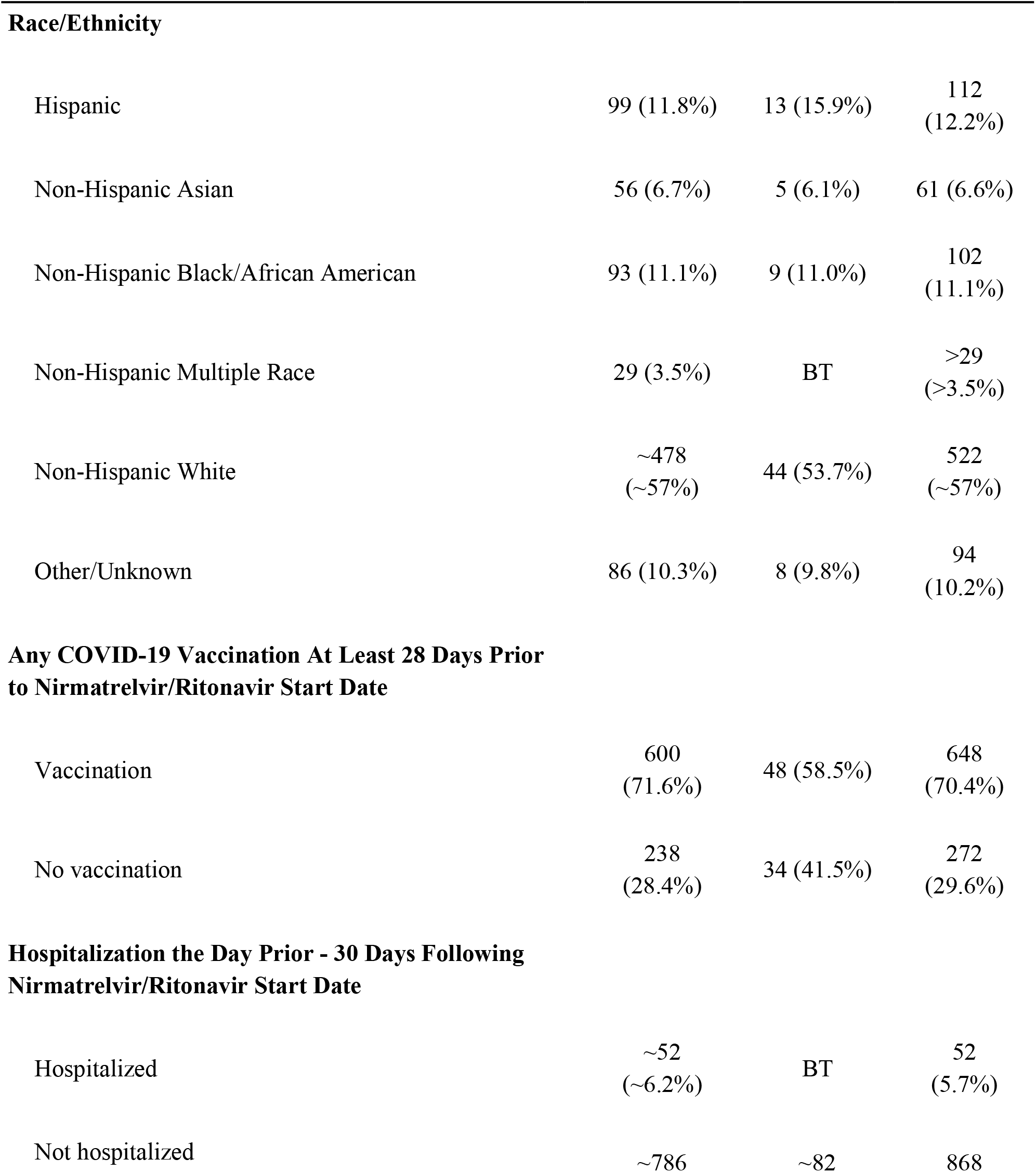
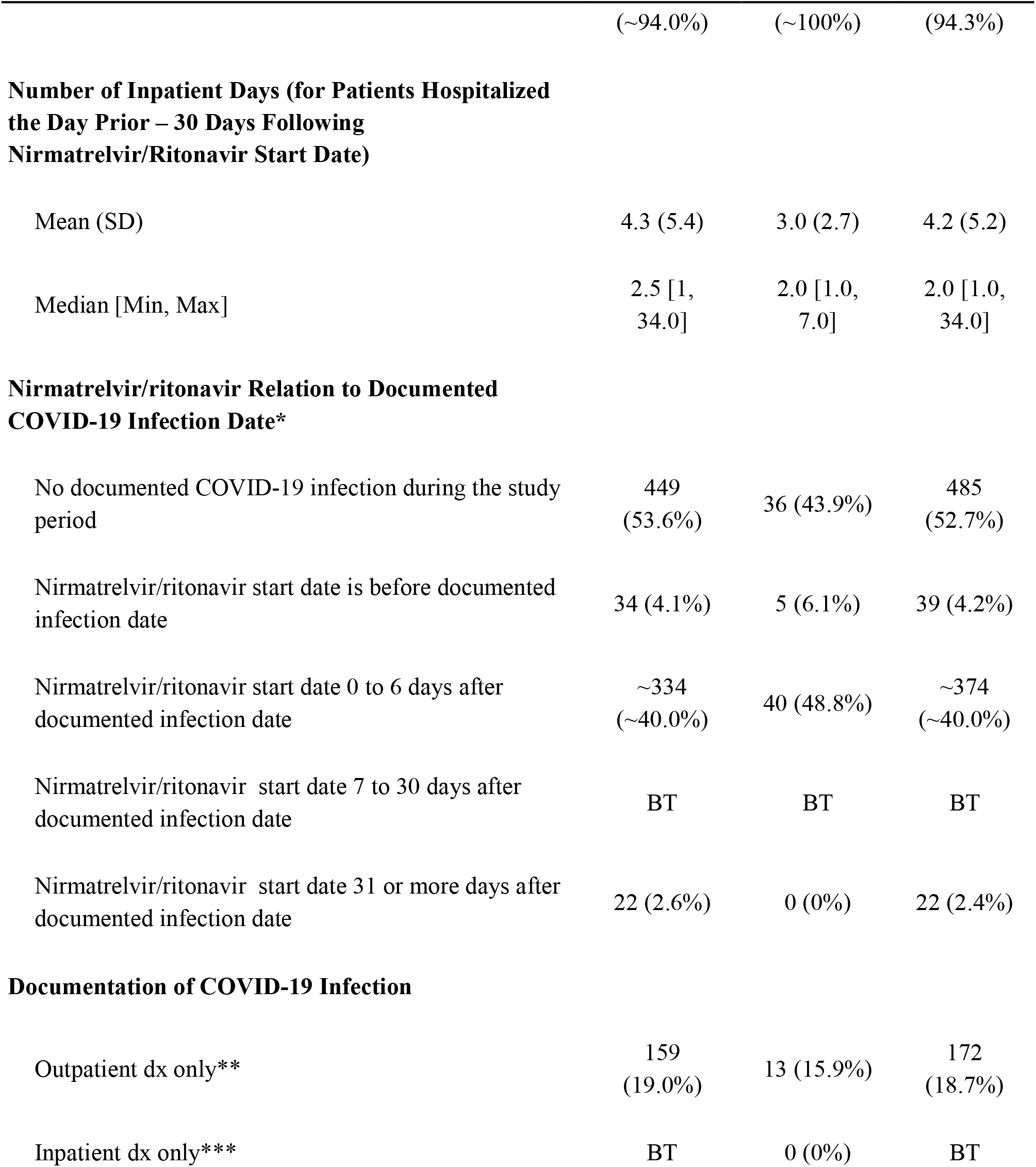
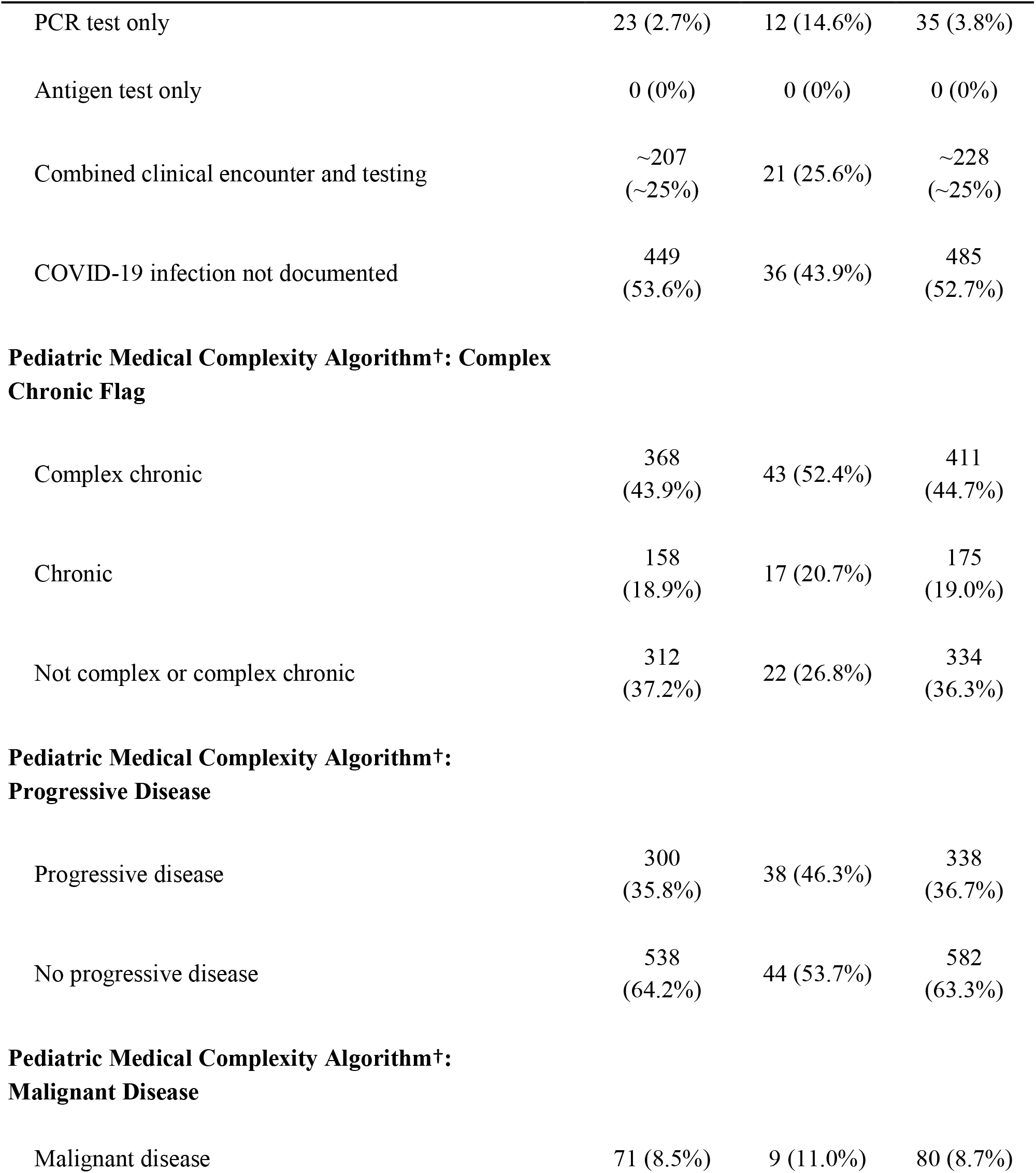
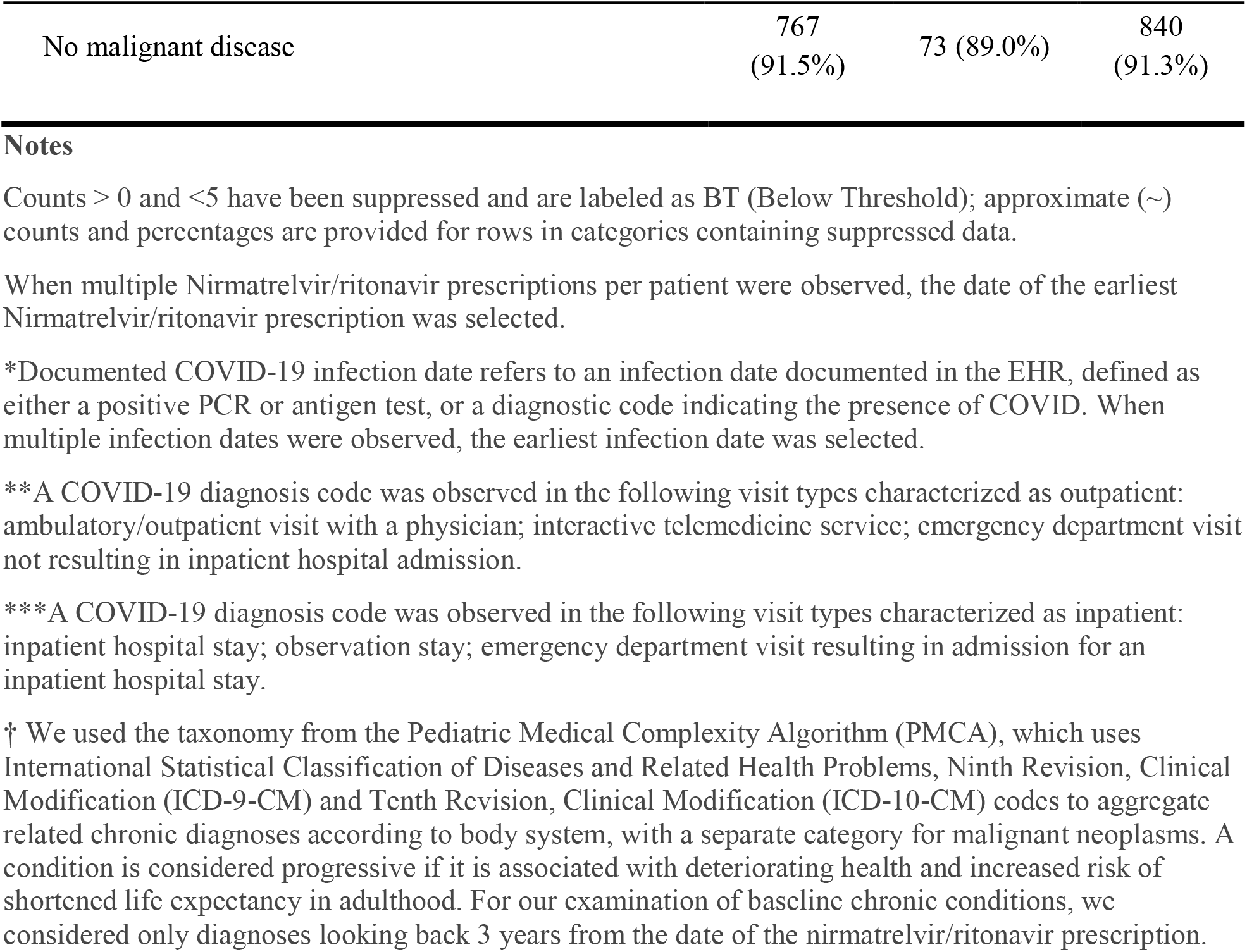
Characteristics of patients receiving nirmatrelvir/ritonavir

**Figure 1.**
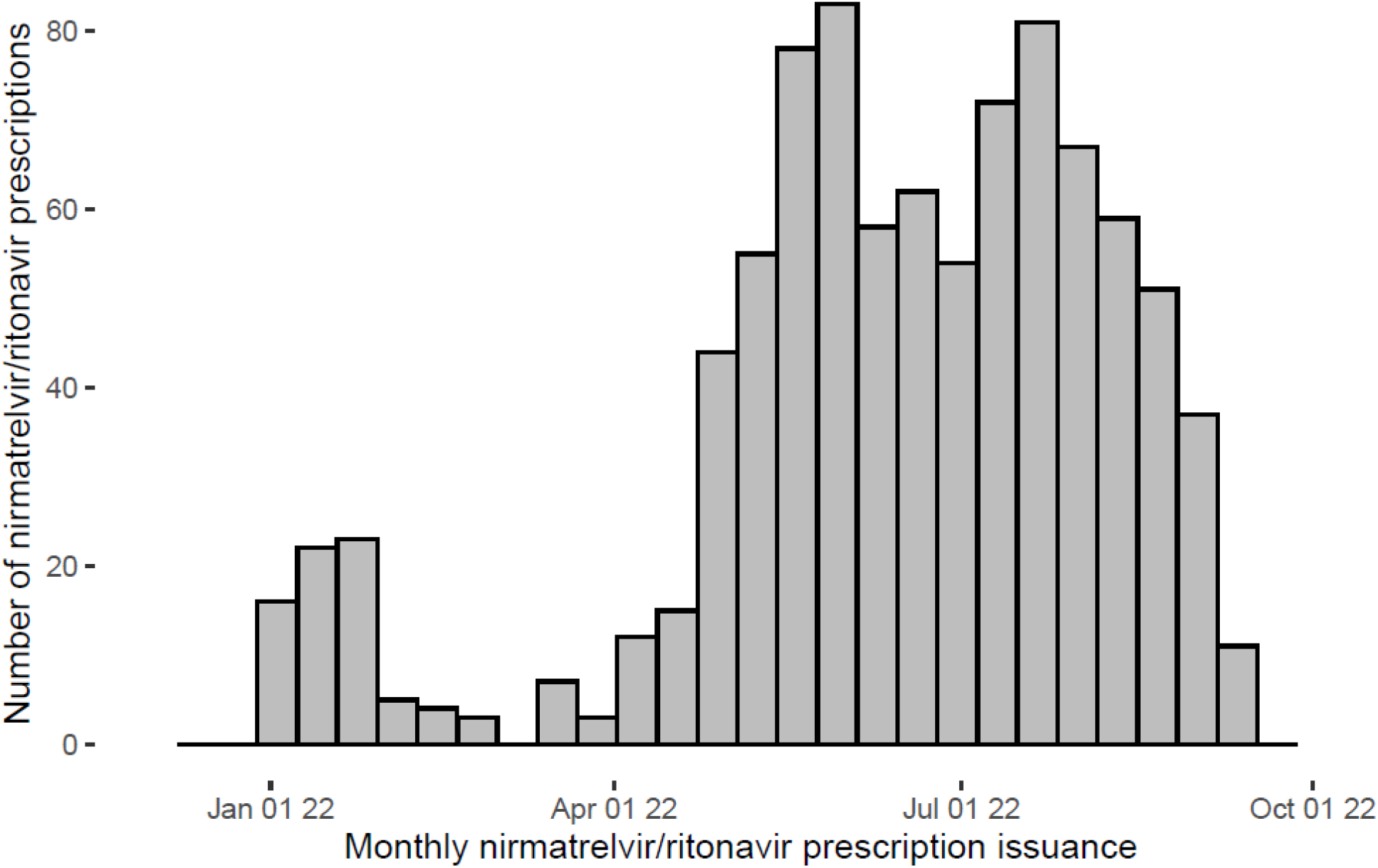
Histogram displaying counts per week of nirmatrelvir/ritonavir prescriptions from December 01, 2021 to September 29, 2022.

Among nirmatrelvir/ritonavir recipients, 64% (586/920) had a chronic or complex chronic disease and 9% (80/920) had a malignant disease.^10^ Only 52/920 (5.7%) of individuals were hospitalized within 30 days following nirmatrelvir/ritonavir prescription.

## DISCUSSION

Nirmatrelvir/ritonavir prescribing within PEDSnet affiliated health systems has steadily increased since April 2022, but remains limited. Within the PEDSnet RECOVER database, a majority of individuals received prescriptions without outpatient encounters or health system-based COVID-19 testing. Factors like ubiquitous home testing for COVID-19 and time-sensitive nirmatrelvir/ritonavir prescribing requirements (within 5 days of symptom onset) may drive the observed paucity of clinical interactions accompanying prescriptions. This limited clinical footprint may hamper efforts to generate pediatric nirmatrelvir/ritonavir evidence through HER data. Strategies to optimize data capture, such as institutional EHR pathways requiring COVID-19 diagnosis association with nirmatrelvir/ritonavir prescribing, can better position learning health networks to evaluate the impact of therapeutics on pediatric COVID-19 outcomes.

Nirmatrelvir/ritonavir recipients had high chronic disease burden, but <6% of recipients were hospitalized within 30 days of receiving a prescription. The diversity of chronic conditions represented within this sample, along with small sample sizes contained within particular disease states, precluded disease specific characterization. While adults aged 18-23 years comprise only 5% of PEDSnet patients^9^, approximately 40% of nirmatrelvir/ritonavir prescriptions observed within this study cohort were issued to individuals in this age group.

This study’s collective findings highlight the urgent need for nirmatrelvir/ritonavir data efficacy and effectiveness data in pediatric chronic disease populations, including comparative effectiveness with other COVID-19 therapeutics. Concurrent investigation to identify disparities in pediatric nirmatrelvir/ritonavir access and use can inform strategies for equitable, evidence-based delivery of COVID-19 therapeutics.

## Data Availability

This retrospective cohort study evaluates electronic health record (EHR) data from 8 PEDSnet health systems. Unlike public data repositories, individual-level datasets are not typically released outside the network. Due to the complexity and privacy concerns associated with electronic health record data, certified PEDSnet data scientists lead the execution of data analyses. For general questions about PEDSnet or the collaboration request process, email the Project Management Office (PMO) at
https://redcap.chop.edu/surveys/?s=88WJ8WAFPJ

## Abbreviations

(U.S.): United States
(FDA): Food and Drug Administration
(EUA): Emergency Use Authorization
(EHR): Electronic health record
(PMCA): Pediatric Medical Complexity Algorithm
(RECOVER): Researching COVID to Enhance Recovery

## Acknowledgments

The authors gratefully acknowledge the contributions of Miranda Higginbotham.

